# Compassionate Use of Tocilizumab in Severe SARS-CoV2 Pneumonia. When late administration is too late

**DOI:** 10.1101/2020.06.13.20130088

**Authors:** Miguel Górgolas Hernández-Mora, Alfonso Cabello Úbeda, Laura Prieto Pérez, Felipe Villar Álvarez, Beatriz Álvarez Álvarez, María Jesús Rodríguez Nieto, Irene Carrillo Acosta, Itziar Fernández Ormaechea, Aws Waleed Mohammed Al-Hayani, Pilar Carballosa, Silvia Calpena Martínez, Farah Ezzine, Marina Castellanos González, Alba Naya, Marta López de las Heras, Marcel José Rodríguez Guzmán, Ana Cordero Guijarro, Antonio Broncano Lavado, Alicia Macías Valcayo, Marta Martín García, Javier Bécares Martínez, Ricardo Fernández Roblas, Miguel Ángel Piris Pinilla, José Fortes Alen, Olga Sánchez Pernaute, Fredeswinda Romero Bueno, Sarah Heili Frades, Germán Peces Barba Romero, the COVID-FJD-TEAM

**Affiliations:** Division of Infectious Diseases, Fundación Jiménez Díaz, Universidad Autónoma de Madrid, Spain; Department of Pneumology, Fundación Jiménez Díaz, Universidad Autónoma de Madrid, Spain; Department of Microbiology, Fundación Jiménez Díaz, Universidad Autónoma de Madrid, Spain; Department of Pharmacy, Fundación Jiménez Díaz; Department of Pathology, Fundación Jiménez Díaz, Universidad Autónoma de Madrid, Spain; Department of Rheumatology, Fundación Jiménez Díaz, Universidad Autónoma de Madrid, Spain

**Keywords:** SARS-CoV-2, Covid-19, Tocilizumab, Pneumonia, Treatment

## Abstract

**Introduction:** Tocilizumab is an interleukin 6 receptor antagonist which has been used for the treatment of severe SARS-CoV-2 pneumonia (SSP), aiming to ameliorate the cytokine release syndrome (CRS) -induced acute respiratory distress syndrome (ARDS). However, there is no data about the best moment for its administration along the course of the disease.

**Methods:** We provided tocilizumab on a compassionate-use basis to patients with SSP hospitalized (excluding intensive care and intubated cases) who required oxygen support to have a saturation >93%. Primary endpoint was intubation or death after 24 hours of its administration. Patients received at least one dose of 400 mg intravenous tocilizumab during March 8-2020, through April 20-2020.

**Findings:** A total of 207 patients were studied and 186 analysed. The mean age was 65 years and 68% were male. A co-existing condition was present in 68 % of cases. At baseline, 114 (61%) required oxygen support with FiO2 >0.5 % and 72 (39%) ≤0.5%. Early administration of tocilizumab, when the need of oxygen support was still below FiO2 ≤0.5%, was significantly more effective than given it in advanced stages (FiO2 >0.5 %), achieving lower rates of intubation or death (13% vs 37% repectively, p<0·001).

**Interpretation:** The benefit of tocilizumab in severe SARS-Cov-2 pneumonia is only expected when it is administrated before the need of high oxygen support.

**Funding:** None.

## Introduction

Since December 2019 the SARS-CoV-2 pandemic has affected more than 6,4 million people worldwide and more than 380,000 fatalities have been recorded [1] at the time of writing. Patients with severe SARS-CoV-2 pneumonia (SSP) die due to poor oxygenation despite ventilatory support and different treatments including drugs with anti-viral activity, such as remdesivir, lopinavir/ritonavir, interferon beta, hydroxychloroquine; and/or anti-inflammatory drugs, such as corticosteroids, azithromycin and low molecular weight heparin amongst other [2–5].

Pathological post-mortem samples of lung and bone marrow of these patients show diffuse alveolar damage with alveolar edema, hyaline membranes and microvascular thrombosis along with extensive hemophacytosis in the bone marrow [6]. Laboratory data show high levels of ferritin, interleukin-6, C-reactive protein, LDH and D dimer, all indicative of a cytokine release syndrome (CRS) - induced ARDS [7–10] derived from the viral infection.

It is believed that the severity of SARS-CoV-2 pneumonia depends not only on the viral load in lung tissue but mainly on the inflammatory response of the host. Interleukin-6 is a key factor for the activation of the cis- and trans-signaling pathways leading to the cytokine release syndrome [11,12]. Tocilizumab is an interleukin 6 receptor antagonist which has been used for the treatment of rheumatoid arthritis [13] and for the treatment of chimeric antigen receptor (CAR) T cell-induced CRS in cancer patients [14–16]. Information about its use for SARS-CoV-2 pneumonia is limited [17,18] and results of randomized clinical trials are still pending [19,20].

We present a cohort of 207 patients treated with tocilizumab at a single institution during the COVID-19 outbreak in Madrid.

## Methods

### Patients

From the 8^th^ of March until 19^th^ of April 2020 a total of 2,416 patients were admitted to the Fundación Jiménez Díaz University Hospital in Madrid, with a confirmed or suspected diagnosis of SARS-CoV-2 pneumonia. This was defined as the presence of unilateral or bilateral lung infiltrates with basal oxygen saturation below 94% in patients with confirmed positive COVID-19 RT-PCR (Viasure® SARS-CoV-2 Real Time PCR detection kit) in nasopharyngeal or throat swabs or, in the absence of microbiological confirmation, the existence of epidemiological data (close contact with documented patients) or laboratory data (lymphopenia, high levels of ferritin, high sensitivity C reactive protein (HSCRP), LDH, interleukin-6, D-dimer) suggestive of COVID-19 infection.

Our standard treatment for SSP included lopinavir/ritonavir (if the disease started fewer than 7 days before admission), hydroxychloroquine or chloroquine, doxycycline or azithromycin, low molecular weight heparin, cyclosporine, n-acetylcysteine, pulse corticosteroids and, in some cases, interferon beta 1-b. This treatment protocol was approved by the Drug Commission of the Fundación Jiménez Díaz University Hospital.

Tocilizumab was recommended if a FiO2 greater than 0.24% was necessary to achieve an oxygen saturation above 93%. A single dose of 400-600 mg of tocilizumab was given intravenously. 17 patients with very severe disease (median FiO2 1%, IQR:0·4-1) received one or two more consecutive doses if the drug was readily available. None of the patients were at the intensive care unit or with high flow oxygen support at the time of tocilizumab administration.

### Study assessments

Data on patient’s oxygen-support at admission, before and after tocilizumab administration were recorded according to standard clinical practice. Laboratory values including absolute lymphocyte counts, serum ferritin, interleukin-6, high sensitive C reactive protein, D-Dimer, serum creatinine, ALT, AST, LDH and lipid profile were also reported as standard clinical practice. The primary endpoint was the need for intubation or death. Patients who required intubation or died within 24 hours after tocilizumab administration were not included in the analysis.

### Program oversight

All patients signed an informed consent for the compassionate use of tocilizumab before its administration. This study was approved by the Medical Ethics Committee of the Fundación Jiménez Díaz University Hospital. All data were collected by the investigators who performed the statistical analysis.

### Statistical analysis

All patients who received at least one dose of tocilizumab between March 8, 2020 until April 20, 2020 were included in the analysis. Distribution normality was assessed using the Kolmogorov-Smirnov test. Normally distributed data were presented as mean (SD), non-normally distributed data as median (IQR), and categorical variables as frequency (%). Differences between groups were analysed by Fisher’s exact test for categorical data or one-way ANOVA for continuous data. Kaplan-Meiers curves were used for survival studies. Results are reported as point estimates and 95 percent confidence intervals. Analysis were done with SPSS software version 24.0.

## Results

### Patients

In total, 207 hospitalized patients received at least one dose of 400 mg iv tocilizumab between March 8, 2020 until April 20, 2020, of whom 21 were excluded of the analysis because were intubated or died in the first 24 hours after tocilizumab administration leaving 186 patients studied. 169 (91%) patients received one dose, 16 patients two doses and 1 patient three doses.

### Baseline characteristics of the patients

The main clinical characteristics of patients are summarized in Table 1. The mean age of patients was 65 years and 68% were male. 68% of patients had a co-existing condition, high blood pressure being the most prevalent (51%). At the time of tocilizumab administration 114 (61%) patients required FiO2 ≥0.5% and 72 (39%) required FiO2 <0.5%. The main laboratory values before tocilizumab administration showed a marked elevation of ferritin, interleukin-6 and C-reactive protein, D-dimer and a low absolute lymphocyte count.

**Table 1.** Clinical Characteristics of Patients with Severe SARS-CoV-2 Pneumonia Treated with Tocilizumab.

|  |  |
| --- | --- |
| **Characteristic** | **N = 186** |
| Mean age (SD)-yr | 65 (11·4) |
| Age category – no. (%) |  |
| < 50 yr | 17 (9·1) |
| 50 to < 70 yr | 97 (52·1) |
| ≥ 70 yr | 72 (38·7) |
| Male sex- no. (%) | 126 (67·7) |
| Ethnicity – no. (%) |  |
| - Caucasian | 177 (95·2) |
| - Latin American | 9 (4·8) |
| - Other | 0 (0) |
| Coexisting conditions – no. (%) |  |
| - None | 59 (31·7) |
| - Hypertension | 94 (50·5) |
| - Diabetes | 39 (21) |
| - Obesity | 57 (30·6) |
| - Vasculopathy | 29 (15·6) |
| - Chronic obstructive lung disease | 13 (7) |
| - Chronic renal failure (GFR < 30 ml/min) | 6 (3·2) |
| - Immunosuppression | 20 (10·8) |
| Median duration of symptoms prior to tocilizumab therapy (IQR) – days | 11 (8-13) |
| Median duration of hospital admission prior to tocilizumab therapy (IQR) – days | 3 (1-5) |

Almost all patients (168p, 90·3%) had received antiretroviral drugs (lopinavir/ritonavir or darunavir/ritonativir) for a median duration of 3 days (IQR:1-5). Hydroxychloroquine or chloroquine sulphate had been administered to 97·8% of cases; cyclosporine to 89·2%, interferon beta-1b to 9·7% and LMWH to 96·2%. Pulse methyl-prednisolone had been given to 95·7% of cases at a dose of 250 mg/day for one to three days before tocilizumab. Antimicrobial agents, either doxicicline or azithromycin or ceftriaxone was given to all patients for a minimum duration of 5 days.

During a follow-up period of fifteen days 51 patients achieved the primary endpoint (intubation or death) 19 patients needed intubation (of whom 4 died) and 36 died (32 of whom were not intubated). The primary endpoint (intubation or death) was significantly different in the group receiving tocilizumab when the oxygen support was high (FiO2 >0.5 %) compare to those with FiO2 ≤0.5% (37% vs 13%, p<0·001) (Figure 1) (Table 2).

**Table 2.** Clinical characteristics of patients with early (FiO2 ≤0.5) and late (FiO2 >0.5) tocilizumab treatment.

|  |  |  |  |
| --- | --- | --- | --- |
| Characteristic | Early (n=72) | Late (n=114) | p |
| Mean age (SD)-yr | 64 (11) | 66 (12) | =0.139 |
| Male sex- no. (%) | 49 (68) | 77 (68) | =0.942 |
| Ethnicity – no. (%) |  |  |  |
| - Caucasian | 68 (94) | 109 (96) | =0.717 |
| - Latin American | 4 (6) | 5 (4) | =0.717 |
| Coexisting conditions – no. (%) |  |  | =0.307 |
| - None | 26 (36) | 33 (29) |  |
| - Hypertension | 34 (47) | 60 (53) | =0.472 |
| - Diabetes | 10 (14) | 29 (25) | =0.059 |
| - Obesity | 15 (21) | 42 (37) | **=0.021** |
| - Vasculopathy | 16 (22) | 13 (11) | **=0.048** |
| - Chronic obstructive lung disease | 7 (10) | 6 (5) | =0.245 |
| - Chronic renal failure (GFR < 30 ml/min) | 4 (6) | 2 (2) | =0.153 |
| - Immunosuppression | 9 (13) | 11 (10) | =0.541 |
| Mean duration of symptoms prior to tocilizumab therapy (SD) – days | 10.7 (5) | 11.7 (6) | =0.237 |
| Mean duration of hospital admission prior to tocilizumab therapy (SD) – days | 3.8 (3) | 3.3 (3) | =0.298 |
| Concomitant treatment – no. (%) |  |  |  |
| - Steroids | 68 (94) | 110 (97) | =0.503 |
| - Protease inhibitors | 62 (86) | 106 (93) | =0.123 |
| - Hydroxychloroquine | 71 (99) | 111 (97) | =0.569 |
| - Cyclosporine | 59 (82) | 107 (94) | **=0.011** |
| - Interferon beta | 5 (7) | 13 (11) | =0.316 |
| - Heparin | 68 (94) | 111 (97) | =0.307 |
| - Antibiotics | 72 (100) | 114 (100) |  |
| Median laboratory values (SD) |  |  |  |
| - Serum ferritin - µg/liter | 1,842 (1,850) | 1,466 (1,443) | =0.139 |
| - High-sensitivity C-reactive protein - mg/dl | 12 (11) | 12 (10) | =0.864 |
| - Interleukin-6 – pg/ml | 156 (329) | 176 (624) | =0.830 |
| - D-dimer - mg/liter | 2,278 (4,709) | 3,864 (13,302) | =0.345 |
| - Absolute lymphocytes - per mm<sup>3</sup> | 862 (630) | 712 (323) | **=0.034** |
| Primary endpoint – no. (%) |  |  |  |
| - Global | 9 (13) | 42 (37) | **<0.000** |
| - Invasive ventilation | 5 (7) | 14 (12) | =0.242 |
| - Death | 4 (6) | 32 (28) | **<0.000** |

**Figure 1.**
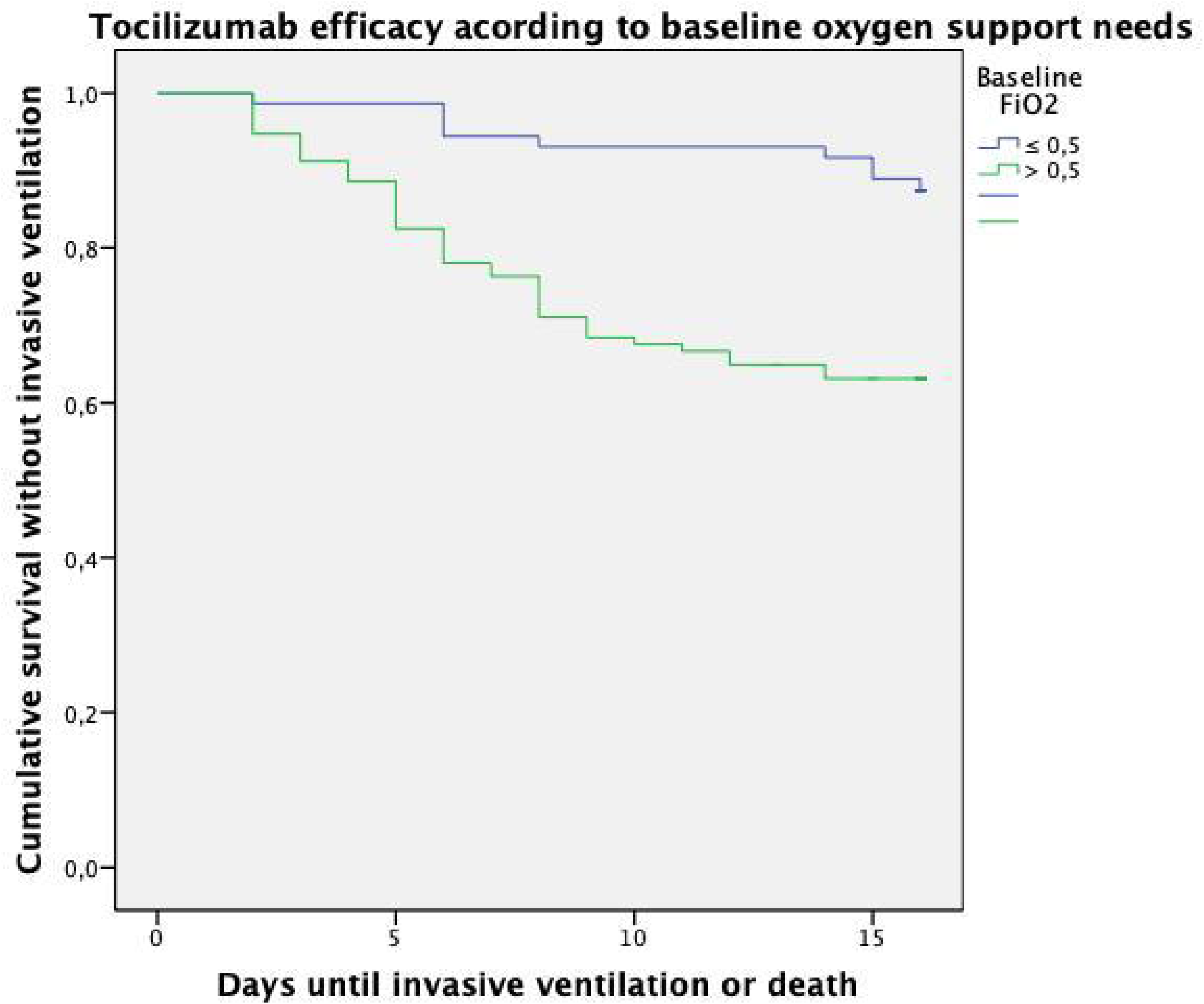
Kaplan-Meiers curves for primary endpoint (need of invasive ventilation or death) in patients treated with tocilizumab. The blue line represents the group of patients who received tocilizumab when their oxygen support needs was low (FiO2 ≤0.5) and the green line represents the group of patients with higher oxygen support needs (FiO2 >0.5) at the time of tocilizumab administration.

Changes in laboratory data after tocilizumab administration are shown in Table 3. A statistically significant decrease in the median serum ferritin and the median HSCRP was observed. Interleukin-6 and D-dimer median serum levels increased and the median absolute lymphocyte count remained stable.

**Table 3.** Clinical, Laboratory, Imaging Data and Outcomes of Patients with Severe SARS-CoV-2 Pneumonia Treated with Tocilizumab.

| Characteristic |  |  |  |
| --- | --- | --- | --- |
|  | Before | After | p |
| Oxygen-support category – no.(%) |  | 7 days after tocilizumab |  |
| - FiO2 – 0.21% (Ambient air) | 2 (1.1) | 38 (20.4) |  |
| - FiO2 – 0.24% | 3 (1.6) | 8 (4.3) |  |
| - FiO2 – 0.28% | 6 (3.2) | 23 (12.3) |  |
| - FiO2 – 0.35% | 36 (19.4) | 22 (11.8) |  |
| - FiO2 – 0.4% | 23 (12.3) | 14 (7.5) |  |
| - FiO2 – 0.6% | 21 (11.3) | 6 (3.2) |  |
| - FiO2 – 1% | 95 (51.1) | 24 (12.9) |  |
| - Intubated/dead |  | 51 (27.4) |  |
| Median laboratory values (IQR) |  |  |  |
| - Serum ferritin - µg/liter | 1,211 (716-2,105) | 1,139 (673-1,880) | =0.003 |
| - High-sensitivity C-reactive protein - mg/dl | 8.9 (3.4-18.9) | 1.4 (0.6-3) | <0.001 |
| - Interleukin-6 – pg/ml | 42 (8.4-100) | 149 (52-519) | <0.001 |
| - D-dimer - mg/liter | 821 (498-1,510) | 1,197 (737-2,240) | <0.001 |
| - Absolute lymphocytes - per mm<sup>3</sup> | 700 (500-900) | 700 (500-1,000) |  |
| Clinical evolution – no. (%) |  |  |  |
| - Endpoint (intubation or death) |  | 51 (27.4) |  |
| - Intubation |  | 19 (10.2) |  |
| - Death |  | 36 (19.4) |  |
| - Live hospital discharge |  | 150 (80.6) |  |
| - Median days from 1<sup>st</sup> dose until discharge |  | 10 (7-15) |  |
| - Median days of hospitalization |  | 14 (7-19) |  |
| Radiological evolution – no. (%) |  |  |  |
| - Improvement |  | 71 (38) |  |
| - Unchanged or deterioration |  | 106 (57) |  |
| - Not available |  | 9 (5) |  |
| Tocilizumab-related serious adverse events – no. (%) |  | 11 (5.9) |  |
| - Increase liver enzymes (AST, ALT) |  | 5 |  |
| - Increase bilirubin |  | 3 |  |
| - Increase creatinin |  | 3 |  |
| - Headache |  | 1 |  |
| - Hyperkalaemia |  | 1 |  |
| Acute infection after tocilizumab – no. (%) |  | 13 (6.9) |  |
| - Fungal |  | 9 |  |
| o *Candida spp* |  | 7** |  |
| o *Aspergillus spp* |  | 2 |  |
| - Bacterial |  | 6 |  |
| o *Pseudomonas aeruginosa* |  | 2 |  |
| o *Klebsiella pneumoniae* |  | 2* |  |
| o *Enterococcus spp* |  | 2* |  |

\*combined with others pathogens

Thirty-six patients died despite tocilizumab treatment. The main demographic, clinical and laboratory data of patients who died and survived is shown in Table 4. Patients who died were older (75·8 years versus 62·5, p<0·001), had any co-existing condition (89% vs 63%, p=0·003), specifically high blood pressure (69% vs 46%, p=0·12); had a higher mean interleukin-6 (1168 vs 311, p<0·001), a higher mean HSCRP (3·5 vs 2·0, p<0·009), a lower absolute lymphocyte count (527 vs 860, p=0·001), and a higher median D-dimer (7832 vs 3190, p=0·027). The global survival rate of those who received tocilizumab was 81% (150p), and it was 94 % for those who received it when their oxygen support was with a FiO2 ≤0.5%, and 72% when it was >0.5% (p=0,000)

**Table 4.** Comparison of the main Characteristics of Patients who Died and Survived after Tocilizumab Treatment.

|  |  |  |  |
| --- | --- | --- | --- |
| **Characteristic** | **Died (n=36)** | **Survived (n=150)** | **p** |
| Mean age – yrs | 75·9 | 62·5 | <0·001 |
| Sex - n° (%) |  |  |  |
| - Male | 24 (67) | 102 (68) | =0·878 |
| - Female | 12 (33) | 48 (32) | =0·878 |
| Any Comorbidity – n° (%) | 32 (89) | 95 (63) | =0·003 |
| HBP – n° (%) | 25 (69) | 69 (46) | =0·012 |
| Diabetes – n° (%) | 10 (28) | 29 (19) | =0·264 |
| Obesity – n° (%) | 13 (36) | 44 (29) | =0·428 |
| Vasculopathy – n° (%) | 9 (25) | 20 (13) | =0·083 |
| Chronic Obstructive Lung Disease – n° (%) | 4 (11) | 9 (6) | =0·280 |
| Cr. Clearance < 30 ml/min – n° (%) | 0 (0) | 6 (4) | =0·223 |
| Immunosuppression – n° (%) | 4 (11) | 16 (11) | =0·938 |
| **Laboratory Data pre and post tocilizumab** |  |  |  |
| Serum ferritin - µg/liter pre | 1,557 | 1,626 | =0·806 |
| Serum ferritin - µg/liter post | 1,366 | 1,338 | =0·898 |
| High-sensitivity C-reactive protein - mg/dl pre | 13·4 | 11·7 | =0·378 |
| High-sensitivity C-reactive protein - mg/dl post | 3·5 | 2·0 | =0·009 |
| Interleukin-6 – pg/ml pre | 389 | 116 | =0·017 |
| Interleukin-6 – pg/ml post | 1,168 | 311 | <0·001 |
| D-dimer - mg/liter pre | 8,288 | 2045 | =0·002 |
| D-dimer - mg/liter post | 7,832 | 3,190 | =0·027 |
| Total lymphocytes - per mm<sup>3</sup> pre | 633 | 803 | =0·052 |
| Total lymphocytes - per mm<sup>3</sup> post | 527 | 860 | =0·001 |

| **Oxygen support pre tocilizumab** |  |  |  |
| --- | --- | --- | --- |
| FiO2 – ≤ 0.5 % pre - n° (%) | 4 (5.6) | 68 (94.4) | <0.001 |
| FiO2 – >0.5 % pre - n° (%) | 32 (28) | 82 (72) | <0.001 |

### Safety

A total of 11 (5·9%) patients had serious adverse reactions related to tocilizumab reported by their treating physicians, including increased hepatic enzymes (5 cases) or bilirubin (3 cases), increased creatinine (3 cases), hyperkalaemia (1 case), and headache (1 case). Secondary acquired infections after tocilizumab administration were documented in 13 cases (6·3%), including fungal (Candida spp 7 cases, Aspergillus spp 2 cases) and bacterial (Pseudomonas aeruginosa 2 cases, Klebsiella pneumoniae 2 cases, Enterococcus spp 2 cases).

## Discussion

SARS-CoV-2 has infected more than 6,4 million people and killed more than 380,000 and, as yet, there is a lack of effective therapy for this novel disease [21]. Several antiviral drugs, such as remdesivir – an RNA polymerase nucleotide analogue - and lopinavir/ritonavir - an HIV protease inhibitor - have been tested either in a limited number of cases or in small clinical trials showing some benefits (remdesivir) [3] or none at all (lopinavir/ritonavir) [2]. However, clinical and pathological studies of SARS-CoV-2 disease indicate that a systemic cytokine storm due to macrophage activation may be the leading cause of death in the vast majority of patients, usually occurring two to four weeks after primary infection [22][14][23]. Therefore, immunomodulatory drugs have been used empirically with the aim of regulating and suppressing the inflammatory reaction that leads to multi-organ failure and death [22][11].

Tocilizumab has previously been tested in a small number of SARS-CoV-2 patients with good results [17]. Our study is, to the best of our knowledge, the largest series presented to date, and gives relevant clinical information about tocilizumab’s efficacy and safety for patients with severe SARS-CoV-2 pneumonia, before randomized clinical trials results are available. Specifically, 150 (81%) out of 186 patients with SSP survived after tocilizumab. Thirty one out of 36 deaths occurred in subjects with co-existing conditions that precluded intubation for mechanical ventilation according to the intensive care committee of the hospital. The most significant result of our study is that early administration of tocilizumab is associated with a good prognosis, avoiding disease progression in 94% of cases, and only 6% requiring intubation due to progressive respiratory insufficiency. In contrast, there is limited benefit when it is administrated in more advanced cases with higher oxygen support needs. Mortality rates in hospitalised patients with SSP varies widely but it is around 15.1% −20% in Spanish studies [24], significantly higher than our series of patient who received tocilizumab early in the course of the disease, when FiO2 requirement was below 0.5%.

Surrogate markers of macrophage activation, such as serum ferritin levels, interleukin-6 levels and high sensitivity C reactive protein changed after tocilizumab therapy, indicating a reduction of the inflammatory process. As expected, the median interleukin-6 levels increased 48 hours after tocilizumab. Of note, the absolute lymphocyte count did not change at all.

Most patients received only one dose of 400 mg tocilizumab, mainly because of shortage of the drug during the peak of the epidemic. Seventeen patients received two or more doses of the drug showing similar efficacy than those who received a single dose. These data suggest that even 400 mg of tocilizumab might be adequate for reducing the acute inflammatory process in this disease.

The safety and tolerance of tocilizumab was good in previous studies of non SARS-CoV-2 patients [24,25]. In our series, a small number of serious adverse events were reported and attributed by physicians to the drug. The acquisition of secondary nosocomial infections was detected in 13 patients (6·9%) being most of them lung or urinary tract infections of fungal or bacterial aetiology. However, all these cases had previously received systemic corticosteroids, cyclosporine and antibiotics, and most of them were admitted in the intensive care unit at the time the secondary infection was detected. This is notable because patients with rheumatoid arthritis or those receiving CAR-T cell therapy for cancer who are treated with long term use of tocilizumab are prone to infectious complications [25][27,28].

In this cohort of 186 patients who received tocilizumab mortality was also statistically related to older age, the presence co-existing medical conditions, particularly a high blood pressure, a low absolute lymphocyte count, a high D-dimer, and higher HSCRP and interleukin-6 serum levels after tocilizumab treatment. These are conditions and surrogate markers that have been previously associated with a poor prognosis.

Our study has several limitations, basically due to the retrospective collection of data and the absence of a control group. Firstly, the decision to administer tocilizumab was made by the medical team responsible for each patient. Therefore, the clinical status of patients and the timing of drug administration after the onset of Covid-19 symptoms were variable; initially it was indicated in very respiratory compromised patients and later it was prescribed much earlier, with lower FiO2 support, letting us study its efficacy in this situation. Secondly, the total amount of drug and number of doses that patients received were not uniform, due, as previously mentioned, to a shortage of the drug in the country during the peak of the epidemic. Thirdly, most patients had received previous and/or concomitant drugs, including systemic corticosteroids and hydroxychloroquine/chloroquine which have anti-inflammatory properties. Therefore, we could not properly assess the impact of these drugs on the overall response of patients treated with tocilizumab. Finally, survival rates would also have been influenced by the UCI committee decision whether a patient was eligible for intubation or not. Only large randomized clinical trials will be able to determine the impact of different immunomodulatory or anti-inflammatory drugs administered simultaneously.

In summary, we present the largest series to date on the compassionate use of tocilizumab in severe SARS-Cov-2 pneumonia, and our data shows that its potential benefit occurs when it is administrated early in the course of the disease, when respiratory compromise is still not very severe, having high survival rates (94%) and very limited side effects and secondary infections.

## Data Availability

All data is included in the manuscript.

## Contributors

MG, AC, LP, OS, FR, SH, and GP conceived and designed the study. LP, IC, OS, and FR contributed to the literature research. FV, BA, MJR, IF, AW, PC, SC, FE, MC, AN, ML, MJR, ACG, AB, AM, MM, and JB contributed to the data collection. RF, MAP, JP, OS, FR, MG, and AC contributed to data interpretation. MG, and AC contributed to the tables elaboration. MG, LP, and GP contributed to the writing of the report.

## Acknowledgments

We would first like to express our deepest gratitude for the patients and their families, who in a time of grief have contributed to the understanding of this disease. We are also grateful to the whole team of physicians and health personnel for their tireless, altruistic dedication, strength and effort during the current pandemics. Finally, we would like to acknowledge Dr. Frances Williams, for her invaluable role as English editor and Laura Cereceda as data manager.

## Appendix

The components’ full names and academics degrees of the COVID-FJD TEAM are as follows:

Belén Arroyo MD, Sonsoles Barrio MD, Marcela Valverde MD, Sheila Recuero MD, Elizabet Petkova MD, Belén Zamarro MD, Mariam Vélez MD, Clara Peiró MD, Soraya de la Fuente MD, Roberto Sierra MD, Javier López Botet MD, Antonio Herranz MD, Jorge Hernández MD, Silvia Rubio MD, Luis Nieto MD, Alicia Estrella MD, Laura Castañeda MD, Jorge Polo Sabau MD, Ana Lucía Rivero Monteagudo MD, Diego Meneses MD, Marta del Palacio Tamarit MD, Elisa Ruiz Arabi MD, Ana Venegas MD, Fernando Tornero Romero MD, Victoria Torrente MD, Pilar Barrio MD, Eduardo Alonso MD, Carolina Dassen MD, Blanca Rodríguez Alonso MD, Myriam Rodríguez Couso MD, Gabriela Rosello MD, Carmen Álvaro MD, Cici Feliz MD, María José Díez Medrano MD, Camila García MD, José Luis Larrea MD, Ana Pello MD, Beatriz González MD, Tatiana Hernández MD, Nancy Sánchez MD, Otto Oliva Oliva MD, Javier Vélez MD, Susana Fraile MD, Maite Ortega MD, Lara Cantero MD, Silvana Scaletti MD, Vanessa Pérez MD, Catalina Martín MD, Teresa Stock MD, Silvia Pérez MD, Andrés Silva MD, Alberto Andrés MD, Marta Oses MD, Miguel Morante MD, Lina Martínez MD, Juliana Botero MD, Diana Fresneda MD, Yolanda Martínez MD, Aida Franganillo MD, Amalia Gil MD, Ana Belén Jiménez MD, Adrián Arapiles MD, María Cruz Aguilera MD, Rafael Rubio MD, Alicia Sánchez MD, Begoña Sánchez MD, Rocío Cardá MD, Jersy Cárdenas MD, Lina Martínez MD, Manuel de la Calle MD, Rafael Touriño MD, José Luis Larrea MD, Miguel Morante MD, Alicia Aurea MD, Marta Monsalvo MD, Iris Martínez MD, Catalina Martín MD, Andrés Silva MD, Blanca Barroso MD, Ana Salomé Pareja MD, Ángel Rodríguez Pérez MD, Raúl Fernández Prado MD, Miguel Ángel Navas MD, Alfonso Romero MD, Ana Nieto Ribeiro MD, Beatriz Giraldez MD, Carolina Gotera Rivera MD, Teresa Gómez García MD, Erwin Javier Pinillos Robles MD, Andrés Giménez Velando MD, Herminia Ortiz Mayoral MD, Francisco José Laso del Hierro MD, Marwan Mohamed Choukri MD, Ainhoa Izquierdo Pérez MD, Laura Núñez García MD, Pablo López Yeste MD, Laura de la Dueña Muñoz MD, Elena Heras Recuero MD, María de los Ángeles Zambrano Chacón MD, Fernanda Troncoso Acevedo MD, Carlos López Chang MD, Elena Cabezas Pastor MD, Abdulkader El Hachem Debek MD, María José Romero Valle MD, Esther Canovas Rodríguez MD, Ángel Miracle MD, Marta González Rodriguez MD, Diana Betancor MD, Nicolás González Mangado MD, Sergio Farrais Villalba MD, Gonzalo Díaz Cano MD, José Manuel Corredera Rodríguez MD, Marina Fernández Ochoa MD, Alicia Gómez-Lopez MD, José María Romero Otero MD, Laura Ortega Martín MD, Leyre Baptista Serna MD, José Antonio Esteban Chapel MD, Andrea Castro-Villacañas Farzamnia MD, Laura Esteban-Lucía MD, Ángel Martínez Pueyo MD, Hans Paul Gaebelt MD.

